# B and T cell immune responses elicited by the BNT162b2 (Pfizer–BioNTech) COVID-19 vaccine in nursing home residents

**DOI:** 10.1101/2021.04.19.21255723

**Authors:** Ignacio Torres, Eliseo Albert, Estela Giménez, María Jesús Alcaraz, Pilar Botija, Paula Amat, María José Remigia, María José Beltrán, Celia Rodado, Dixie Huntley, Beatriz Olea, David Navarro

## Abstract

**Objectives:** The immunogenicity of the BNT162b2 COVID-19 vaccine is understudied in elderly people with comorbidities. We assessed SARS-CoV-2-S-targeted antibody and T cell responses following full vaccination in nursing home residents (NHR).

**Methods:** We recruited 60 NHR (44 female; median age, 87.5 years), of whom 10 had previously had COVID-19, and 18 healthy controls (15 female; median age, 48.5 years). Pre- and post-vaccination blood specimens were available for quantitation of total antibodies binding RBD and enumeration of SARS-CoV-2-S-reactive IFN-γ CD4^+^ and CD8^+^ T cells by flow cytometry.

**Results:** The seroconversion rate in presumably SARS-CoV-2 naïve NHR (95.3%), either with or without comorbidities, was similar to controls (94.4%). A robust booster effect was documented in NHR with prior COVID-19. Plasma antibody levels were higher in convalescent NHR than in individuals across the other two groups. A large percentage of NHR had SARS-CoV-2 S-reactive IFN-γ CD8^+^ and/or CD4^+^ T cells at baseline, in contrast to healthy controls. Either CD8^+^ and/or CD4^+^ T-cell responses were documented in all control subjects after vaccination. Contrariwise, the percentage of NHR exhibiting detectable SARS-CoV-2 IFN-γ CD8^+^ or CD4^+^ T-cell responses (or both), irrespective of their baseline SARS-CoV-2 infection status, dropped consistently after vaccination. Overall, SARS-CoV-2 IFN-γ CD8^+^ and CD4^+^ T-cell responses in NHR decreased in post-vaccination specimens.

**Conclusion:** The BNT162b2 COVID-19 vaccine elicits robust SARS-CoV-2-S antibody responses in NHR. Nevertheless, the frequency and magnitude of detectable SARS-CoV-2 IFN-γ T-cell responses after vaccination was lower in NHR compared to controls.

## INTRODUCTION

The BNT162b2 (Pfizer–BioNTech) COVID-19 vaccine, a nucleoside-modified messenger RNA that encodes the full-length transmembrane S glycoprotein locked in its perfusion conformation, elicits high levels of serum neutralizing antibodies (NtAb), mainly targeting the SARS-CoV-2 receptor-binding domain (RBD), and strong TH_1_-skewed functional CD4^+^ and CD8^+^ T cell responses in experimental models and humans [1–4]. The efficacy of the vaccine has been shown to approach 95% in preventing severe COVID-19 across a wide range of age groups [5]. Nevertheless, there is scarce information as to the immunogenicity and efficacy of this vaccine in elderly people with comorbidities and frailty [6–7], who have been prioritized for vaccination worldwide due to their increased risk of developing severe clinical forms of COVID-19 [8]; indeed, this subset was underrepresented in a phase III clinical trial [5]. In this regard, data from the BNT162b2 COVID-19 vaccine phase I trial suggested reduced lower antibody responses in older people compared to younger participants [2]. To gain further insight into this issue, here we assessed SARS-CoV-2-S targeted antibody and functional T cell responses after vaccination with BNT162b2 in a cohort of nursing home residents (NHR), most displaying one or more comorbidities, either presumably SARS-CoV-2 naïve or with documented prior SARS-CoV-2 infection.

## MATERIAL AND METHODS

### Participants and study design

We enrolled a total of 60 subjects (44 female) onto the study, randomly selected from two NH affiliated to the Clínico-Malvarrosa Health Department, Valencia (Spain), which together provide care for 226 residents. The median age of participants was 87.5 years (range, 53-100). As shown in Table 1, 51 subjects (84%) had one or more comorbidities at enrollment (median, 4; range, 1-7). A total of 18 healthy individuals (15 female) aged a median of 48.5 years (range, 27 to 60 years) with no history of SARS-CoV-2 infection at baseline served as controls. Baseline blood specimens were collected within one week before first vaccine dose (from January 2021 to mid February 2021) in both NHR and controls. Post-vaccination specimens were drawn at a median of 17. 5 days (range, 14-35 days) or 15 days (range, 13-35 days) after the second dose in NHR and controls, respectively (from February 2021 to mid March 2021). Blood specimens from participants were collected in sodium heparin tubes (Beckton Dickinson, U.K. Ltd., UK). Plasma specimens were separated following centrifugation and cryopreserved at −20 °C. Informed consent was obtained from participants. The study was approved by the Hospital Clínico Universitario INCLIVA Research Ethics Committee (February, 2021).

**Table 1.** **Baseline characteristics of nursing home residents included in the study**

| **Characteristic** | **Nursing home residents**<br>**no. (%)** |
| --- | --- |
| **Sex** |  |
| Male | 16 (27) |
| Female | 44 (73) |
| **Comorbidities ($\geq 1$)** | 51 (84) |
| Hypertension | 40 (77) |
| Dyslipidemia | 34 (65) |
| Chronic heart disease | 18 (35) |
| Diabetes mellitus | 16 (31) |
| Vascular disease | 14 (27) |
| Cancer | 10 (19) |
| Chronic renal disease | 10 (19) |
| Chronic respiratory<br>disease | 9 (17) |
| Hyperuricemia | 7 (13) |
| Obesity | 2 (4) |

### Immunogenicity assessment

#### Antibody assays

The following immunoassays were used in the current study: (i) Roche Elecsys® Anti-SARS-CoV-2 S (Roche Diagnostics, Pleasanton, CA, USA), an electrochemiluminescence sandwich immunoassay (ECLIA) that quantifies total (IgG and IgM) antibodies directed against RBD. The assay is calibrated with the first WHO International Standard and Reference Panel for anti-SARS-CoV-2 antibody [9]. The limit of detection of the assay is 0.4 U/ml and its quantification range is between 0.8 and 250 U/mL; (ii) Elecsys® Anti-SARS-CoV-2 (Roche Diagnostics) a qualitative ECLIA detecting IgG and IgM antibodies against SARS-CoV-2 nucleoprotein. Both assays were run on cobas® e601 modular analyzer (Roche Diagnostics, Rotkreuz, Switzerland). Plasma specimens were further diluted (1/10) for antibody quantitation when appropriate. (iii) LIAISON® SARS-CoV-2 TrimericS IgG assay (Diasorin S.p.A, Saluggia, Italy), run on a DiaSorin LIAISON platform (DiaSorin, Stillwater, USA), which measures IgG antibodies against a trimeric S-protein antigen. Samples yielding <13 AU/ml were considered negative. Immunoassays were performed and interpreted following the instructions of the respective manufacturers. Cryopreserved plasma specimens were thawed and assayed in singlets within one month after collection. Baseline and follow-up specimens from a given participant were analyzed in the same run.

#### T cell immunity assay

SARS□CoV□2□reactive IFNγ□producing□CD8^+^ and CD4^+^ T cells were enumerated by flow cytometry for ICS (BD Fastimmune, BD□Beckton Dickinson and Company□Biosciences, San Jose, CA), as previously described [10,11]. Briefly, heparinized whole blood (0.5□mL) was simultaneously stimulated for 6□h with two sets of 15□mer overlapping peptides (11□mer overlap) encompassing the SARS□CoV□2 Spike (S) glycoprotein (S1, 158 peptides and S2, 157 peptides) at a concentration of 1□μg/mL per peptide, in the presence of 1□μg/ml of costimulatory monoclonal antibodies (mAbs) to CD28 and CD49d. Peptide mixes were obtained from JPT peptide Technologies GmbH (Berlin, Germany). Samples mock-stimulated with phosphate□buffered saline (PBS)/dimethyl sulfoxide and costimulatory antibodies were run in parallel. Brefeldin A (10□μg/mL) was added for the last 4□h of incubation. Blood was then lysed (BD FACS lysing solution) and frozen at −80°C until tested. On the day of testing, stimulated blood was thawed at 37°C, washed, permeabilized (BD permeabilizing solution) and stained with a combination of labeled mAbs (anti□IFNγ□FITC, anti□CD4□PE, anti□CD8□PerCP□Cy5.5, and anti□CD3□APC) for 1□h at room temperature. Appropriate positive (phytohemagglutinin) and isotype controls were used. Cells were then washed, resuspended in 200□μL of 1% paraformaldehyde in PBS, and analyzed within 2□h on an FACSCanto flow cytometer using DIVA v8 software (BD Biosciences Immunocytometry Systems, San Jose, CA). CD3^+^/CD8^+^ or CD3^+^/CD4^+^ events were gated and then analyzed for IFN□γ production. All data were corrected for background IFN-γ production and expressed as a percentage of total CD8^+^ or CD4^+^ T cells. Representative flow cytometry plots are shown in Supplementary Figure 1.

#### Statistical methods

Frequency comparisons for categorical variables were carried out using the Fisher exact test. Differences between medians were compared using the Mann–Whitney U-test or the Wilcoxon test for unpaired and paired data, when appropriate. Two-sided exact *P*-values were reported. A *P*-value <0.05 was considered statistically significant. The analyses were performed using SPSS version 20.0 (SPSS, Chicago, IL, USA).

## RESULTS

### SARS-CoV-2-specific antibodies in NHR and controls

No serological evidence of prior SARS-CoV-2 infection was found in 49 (83%) of the 59 NHR at baseline. Pre-vaccination plasma was not available from one patient. In addition, these subjects had been tested at least once for presence of SARS-CoV-2 RNA in nasopharyngeal specimens since the beginning of the epidemic, as a part of a local public health policy for nursing homes, systematically returning negative results. Ten (17%) NHR had suffered from COVID-19, as evidenced by presence of SARS-CoV-2 S and N-specific antibodies and a history of compatible clinical picture and one or more RT-PCR positive results in nasopharyngeal specimens (convalescent NHR).

Plasma collected after the second vaccine dose was available for 43 of the 49 NHR with no documented prior infection. One of the remaining six patients died before having received the second dose. Forty-one out of the 43 subjects tested positive by Roche SARS-CoV-2-S immunoassay. All but one specimen tested negative by SARS-CoV-2 N immunoassay, suggesting that one NHR had presumably contracted SARS-CoV-2 infection between the first and the second vaccine dose. Therefore, the overall seroconversion rate in this NHR group was 95.3%. Of interest, plasma specimens from NHR and controls testing negative were run with LIAISON® SARS-CoV-2 TrimericS IgG assay, also returning negative results. All 10 convalescent NHR had detectable SARS-CoV-2 S and N-binding antibodies at baseline; one patient died before receiving the second vaccine dose. A booster effect was observed in all nine individuals following full dose vaccination, with a median 33-fold (range, 10 to 600-fold) increase in antibody levels. Seventeen out of 18 controls had SARS-CoV-2 S-binding antibodies after the second vaccine dose, while none tested positive for N-specific antibodies. Accordingly, the seroconversion rate in this subgroup was 94.4%. As shown in Figure 1, plasma levels of SARS-CoV-2 S antibodies following complete vaccination were higher (*P*<0.01) in convalescent patients (all 2,500 IU/ml) than in presumably SARS-CoV-2 naïve NHR (median 1120 IU/ml; range, 1.08-2,500) or controls (median 2,211 IU/ml; 168 range, 18.4-2,500).

**Figure 1.**
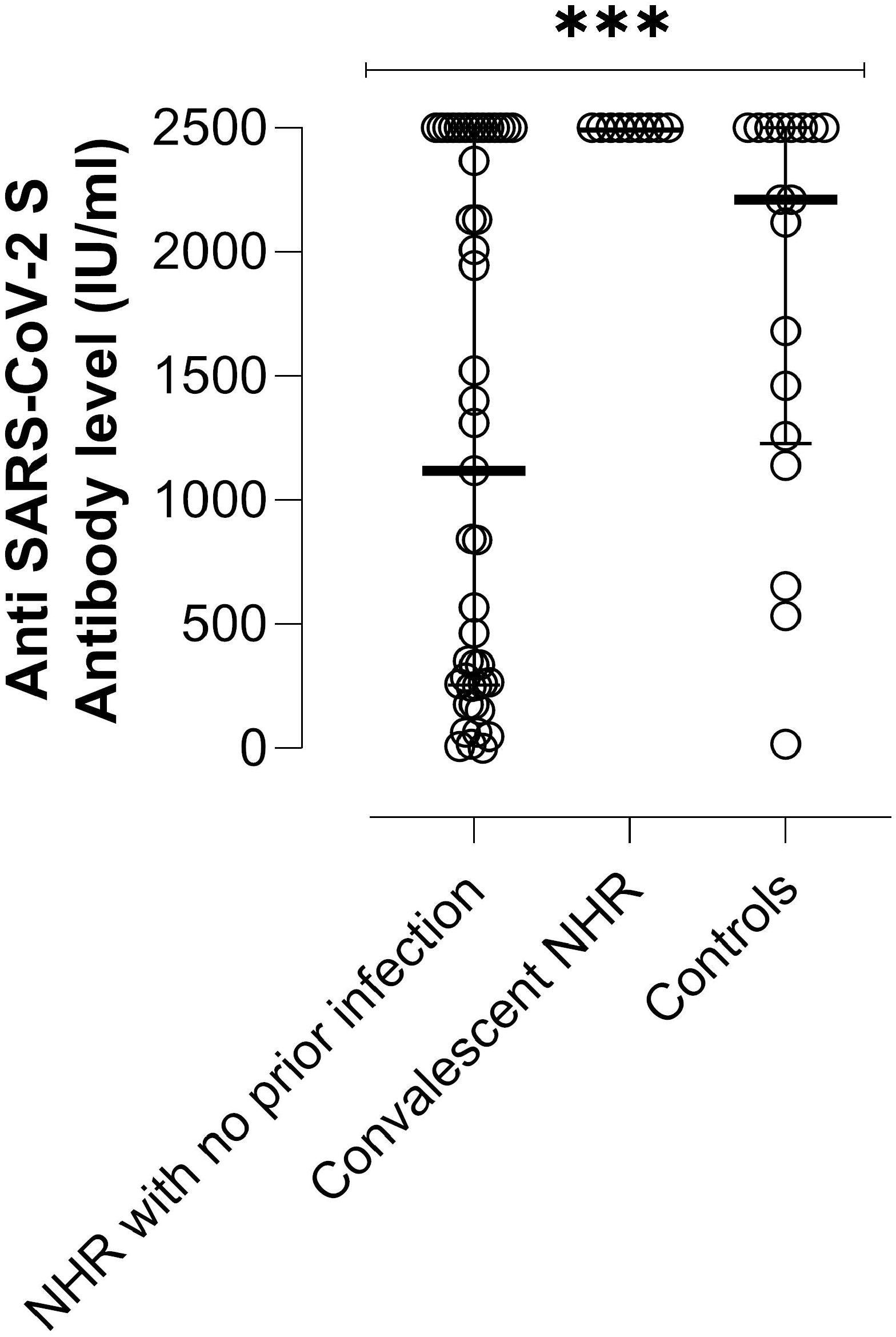
SARS-CoV-2-S plasma antibody levels as measured by Roche Elecsys® Anti-SARS-CoV-2 S immunoassay in nursing home residents (NHR) with or without documented prior SARS-CoV-2 infection and healthy controls following complete vaccination. The asterisks indicate a significant difference in antibody levels across 373 groups (*P*<0.01).

Among NHR with no documented prior infection, the seroconversion rate was comparable (*P*>0.99) in individuals presenting either with (33/35; 94%) or without comorbidities (9/9; 100%). Moreover, having a comorbidity did not impact significantly (*P*=0.14) on SARS-CoV-2-S antibody levels in this population group (Supplementary Figure 2).

### SARS-CoV-2-S-specific T cells in NHR and controls

Analysis of pre-vaccination blood specimens revealed the presence of SARS-CoV-2 S-reactive IFN-γ CD8^+^ and CD4^+^ T cells in 18 (36.7%) and 30 (61.2%) of 49 naïve NHR, and in 6 (60%) and 8 (80%) out of 10 infected NHR; these figures were substantially lower in healthy controls (17.6% for CD8^+^ and 29% for CD4^+^ T cells) (Table 2). Following the second vaccine dose, all control subjects had detectable SARS-CoV-2 S-reactive IFN-γ CD4^+^ T cells and 88% had both IFN-γ CD4^+^ and CD8^+^ T cells. Conversely, the percentage of NHR exhibiting detectable SARS-CoV-2 IFN-γ CD8^+^ or CD4^+^ T cell responses (or both), independently of their baseline SARS-CoV-2 infection status, dropped consistently after vaccination (except for CD8^+^ T cells in NHR without prior infection), as shown in Table 2. Both loss and *de novo* acquisition of detectable SARS-CoV-2 IFN-γ CD8^+^ or CD4^+^ T-cell responses were observed in some individuals, particularly in CD8^+^ T cells. Overall, the magnitude of SARS-CoV-2 IFN-γ CD8^+^ or CD4^+^ T cell responses in NHR, irrespective of their SARS-CoV.2 infection status, decreased consistently in post-vaccination specimens, as can be seen in Table 3. The opposite was observed for healthy controls. The same differential kinetics pattern between NHR and controls was noticed when individuals with detectable T cell responses at baseline were analyzed separately (Figure 2).

**Table 2.** **Detection of SARS-CoV-2-S-reactive T cells in pre- and post-vaccination blood specimens from nursing home residents and controls**

|  |  |  |  |
| --- | --- | --- | --- |
| **Study group** | **SARS-CoV-2-S IFN-γ-producing T cells in pre/post-vaccination peripheral blood specimens** |  |  |
|  | No. of subjects with detectable CD8<sup>+</sup> T-cell response (%) | No. of subjects with detectable CD4<sup>+</sup> T-cell response | No. of subjects with detectable CD8<sup>+</sup> and CD4<sup>+</sup> T-cell responses |
| Nursing home residents with no documented prior SARS-CoV-2 infection | 18 (36) / 27 (54) | 30 (61) / 23 (46) | 17 (34) / 13 (26) |
| Nursing home residents with prior SARS-CoV-2 infection | 6 (60) / 5 (50) | 8 (80) / 7 (70) | 6 (60) / 4 (40) |
| Healthy controls | 3 (17.6) / 15 (88) | 5 (29) / 17 (100) | 1 (6) / 15 (88) |

**Table 3.**
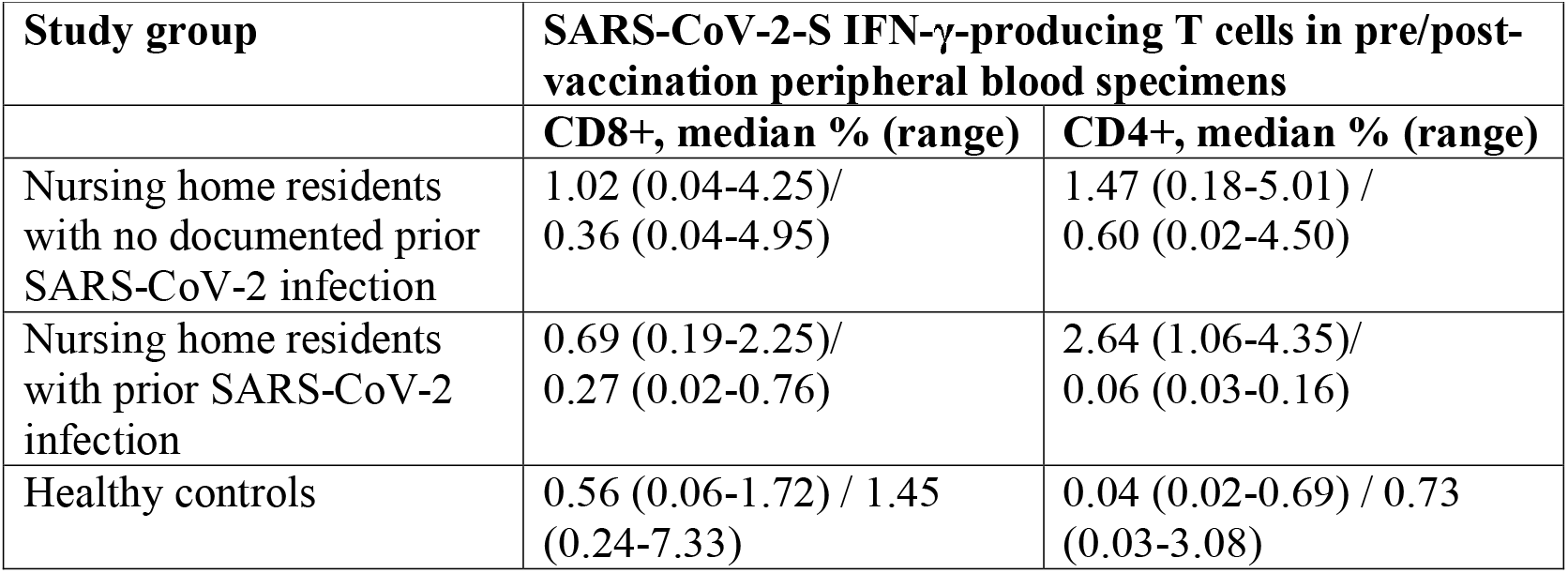
**Enumeration of SARS-CoV-2-S-reactive T cells in pre- and post-vaccination peripheral blood specimens from nursing home residents and healthy controls**

| Study group | SARS-CoV-2-S IFN-γ-producing T cells in pre/post-vaccination peripheral blood specimens | |
| --- | --- | --- |
|  | CD8+, median % (range) | CD4+, median % (range) |
| Nursing home residents with no documented prior SARS-CoV-2 infection | 1.02 (0.04-4.25)/<br>0.36 (0.04-4.95) | 1.47 (0.18-5.01) /<br>0.60 (0.02-4.50) |
| Nursing home residents with prior SARS-CoV-2 infection | 0.69 (0.19-2.25)/<br>0.27 (0.02-0.76) | 2.64 (1.06-4.35)/<br>0.06 (0.03-0.16) |
| Healthy controls | 0.56 (0.06-1.72) / 1.45<br>(0.24-7.33) | 0.04 (0.02-0.69) / 0.73<br>(0.03-3.08) |

**Figure 2.**
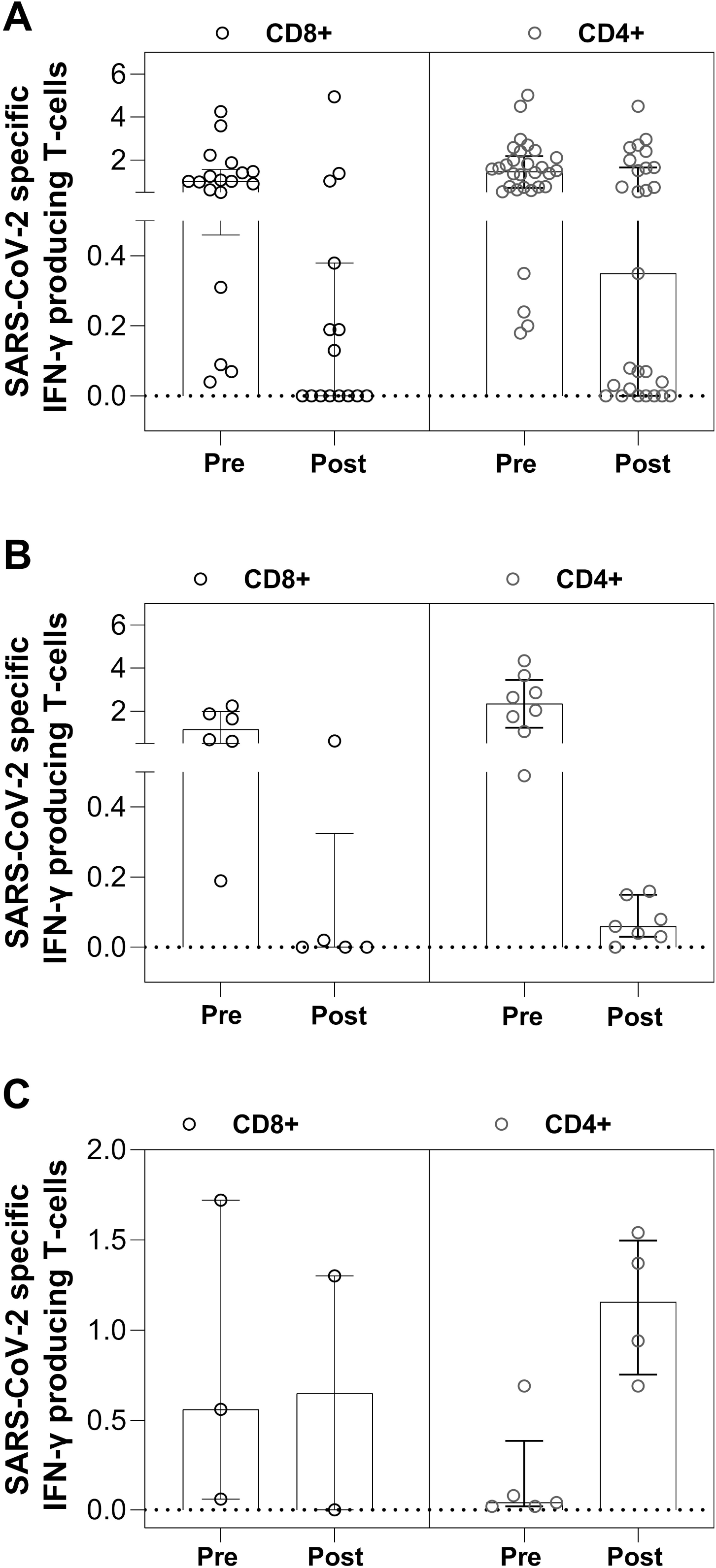
Pre- and post-vaccination SARS-CoV-2-S-reactive IFN-γ-producing CD8^+^ or CD4^+^ T-cell levels in presumably SASR-CoV-2-naïve nursing home residents (NHR) (A), NHR with prior documented SARS-CoV-2 infection (B) and controls (C) with detectable CD8^+^, CD4^+^ T-cell responses or both at baseline.

Overall, the rate and magnitude of detectable SARS-CoV-2 S-reactive IFN-γ CD8^+^ and CD4^+^ T cell responses following vaccination were comparable in NHR with or without comorbidities (Supplementary Table 1).

## DISCUSSION

Data on the immunogenicity of the BNT162b2 COVID-19 vaccine in elderly people with comorbidities is relatively limited. The phase I trial [2] included 12 participants aged 55-85 years and assessed only B cell immunity using a SARS-CoV-2 serum neutralization assay and RBD or S1-binding antibody assays. Slightly weaker antibody responses were reported in older people than in younger participants [2]. Brockman et al. [6] found suboptimal antibody responses after the first vaccine dose in long-term care facility residents as compared to controls. Collier et al. [7] evaluated SARS-CoV-2-S-targeted B and T cell responses elicited by multiplex particle-based flow cytometry, and T cell responses as measured by a CD3^+^ IFN-γ Fluorospot, in 50 participants with a median age of 81 years. They found comparable serum antibody neutralization titers and SARS-CoV-2 S-reactive T cell numbers in participants 80 years or above as in those under 80 following the second vaccine dose.

Here, we quantified total antibodies binding SARS-CoV-2 RBD by means of an ECLIA normalized to the first WHO international standard [9], which strongly correlate with neutralizing antibody titers [12,13]. In turn, SARS-CoV-2-S-reactive IFN-γ-producing CD8^+^ and CD4^+^ T cell were enumerated using a whole blood flow cytometry assay [10,11] at a median of 2-3 weeks after the second vaccine dose. Most NHR recruited (median age, 87.5 years) had one or more comorbidities (84%), and were either with or without a SARS-CoV-2 diagnosis by serological and molecular assay prior to vaccination. The main findings of the study are summarized as follows.

First, overall, the SARS-CoV-2-S seroconversion rate was similar in NHR (95.2%) and controls (94.4%), with no significant differences in median antibody levels across groups. To rule out the presence of antibodies targeting epitopes outside RBD in plasma specimens from NHR and controls testing negative, these were run with a SARS-CoV-2 TrimericS IgG assay, which also returned negative results. Second, a dramatic booster effect was documented in all NHR previously infected by SARS-CoV-2; in fact, these subjects reached significantly higher antibody levels than those measured in presumably naïve NHR and controls.

Third, while detectable SARS-CoV-2 S-reactive IFN-γ CD8^+^ and/or CD4^+^ T-cell responses were documented in post-vaccination specimens from all control subjects, they were present in 50%-70% of NHR, depending upon the T cell subset considered and whether or not subjects had prior history of SARS-CoV-2 infection. In this context, NHR individuals with no prior documentation of SARS-CoV-2 infection appeared to display poorer post-vaccination T cell responses than convalescent NHR. Moreover, in contrast to controls, a consistent decrease in the number of SARS-CoV-2-S-reactive IFN-γ CD8^+^ and CD4^+^ T cells was noticed in post-vaccination specimens from most NHR, regardless of their SARS-CoV-2 infection status. Interpreting the T cell response data presented herein is shadowed by difficulty in ascertaining the true infection status of NHR and controls, regarding which a differential effect of the second BNT162b2 dose on T and B cell immunity was reported in COVID-19-naïve and recovered individuals, with the latter exhibiting poorer responses [14,15]. In effect, functional SARS-CoV-2-S IFN-γ T cells detected in pre-vaccination specimens from NHR and controls with no evidence of prior SARS-CoV-2 infection may well have been seasonal coronavirus cross-reactive T cells, reported to be present in up to 60% (for CD4^+^ T cells) of pre-pandemic blood specimens [see 16 for review]. Circulation of seasonal coronaviruses in NH facilities and repeat exposure of residents is common over the winter season. This may account for the large percentage of NHR with no documented prior SARS-CoV-2 infection displaying T cell responses at baseline (60% for CD4^+^ T cells). Nevertheless, we cannot rule out that some of the current study participants, NHR in particular, could have been asymptomatically infected and failed to mount durable antibody responses, despite robustly expanding SARS-CoV-2-specific T cells [17]. This is plausible, as several SARS-CoV-2 outbreaks were declared in both NH during 2020. Likewise, healthy controls in this study were laboratory employees or staff at the Microbiology unit, and could have been exposed and infected. Regardless of the true SARS-CoV-2 infection status of participants, NHR displayed poorer SARS-CoV-2 T-cell responses than healthy controls after vaccination. In contrast, Collier et al. [7] found no age-related differences in T-cell response after full vaccination dose, although the authors admitted they were unable to adjust for confounders such as immune suppression and comorbidities and had no information on pre-vaccination SARS-CoV-2 infection status of participants. Our findings could be partly explained by the detrimental impact of age-related immunosenescence on immune response to vaccines [18]. Nonetheless, there is biological and clinical use in elucidating whether either cross-reactive or truly specific pre-existing immunity to SARS-CoV-2 may qualitatively or quantitatively modulate vaccine-elicited T cell immune responses, and if so, how this translates into effective protection against the virus. The apparent contraction of SARS-CoV-2-S-reactive IFN-γ T cells in convalescent NHR following the second vaccine dose was also observed by Camara et al. [14], who hypothesized that this second dose may functionally exhaust SARS-CoV-2-S-specific T cells. This may also apply to cross-reactive T cells. In this sense, CD4^+^ T cell responses against common cold coronaviruses (CCC) were decreased in SARS-CoV-2-infected health care workers, suggesting that exposure to SARS-CoV-2 might somehow interfere with CCC responses [19]. Whether this might also be the case following vaccination needs to be defined. Fourth, comorbidities did not appear to have a major impact on either seroconversion rate or magnitude of antibody or T-cell responses following the second vaccine dose in NHR.

The current study has several limitations that must be underlined. Firstly, the number of participants was relatively limited; second, the possibility that NHR displayed SARS-CoV-2-S-reactive T cells with functional specificities other than IFN-γ production was not explored. Additionally, a whole-blood flow cytometry assay was used to assess T-cell immunity: it is uncertain whether employing isolated peripheral blood mononuclear cells instead would increase sensitivity. Finally, no attempt was made to differentiate between truly SARS-CoV-2-specific and cross-reactive T cells, which can be accomplished according to recent reports, although this need further validation [20,21].

In summary, we were able to document robust SARS-CoV-2-S antibody responses equivalent to those of healthy controls in NHR following complete vaccination, irrespective of SARS-CoV-2 infection status and presence or absence of comorbidities. Nevertheless, our data point to differential vaccine effectivity between NHR and controls in terms of eliciting SARS-CoV-2 IFN-γ T-cell responses. In this context, the potential detrimental effect of pre-existing *bona fide* or cross-reactive SARS-CoV-2 immunity seen in NHR merits further investigation.

## Data Availability

The data that support the findings of this study are available on request from the corresponding author, DN.

## ACKNOWLEDGMENTS

We are grateful to all personnel who work at NHR affiliated to the Health Department Clínico-Malvarrosa and at Clinic University Hospital, in particular those at Microbiology laboratory, for their commitment in the fight against COVID-19. Eliseo Albert holds a Juan Rodés Contract (JR20/00011) from the Health Institute Carlos III. Ignacio Torres holds a Río Hortega Contract (CM20/00090) the Health Institute Carlos III.

## AUTHOR CONTRIBUTIONS

EA, EG, MJA, PA, MJR, IT, DH and BO: Methodology and data validation. PB, MJB, R: in charge of implementing public health policies to combat SARS-CoV-2 epidemic at NHR affiliated to the Health Department Clínico-Malvarrosa. DN: Conceptualization, supervision, writing the original draft. All authors reviewed the original draft.

## FIGURE LEGENDS

**Supplementary Figure 1**. Flow cytometry plots corresponding to representative examples of individuals either with or without detectable SARS-CoV-2-S-reactive IFN-γ-producing CD8+ (upper panels) or CD4^+^ (lower panels) T cells. Heparinized whole blood was simultaneously stimulated with two sets of 15□mer overlapping peptides (11□mer overlap) encompassing the SARS□CoV□2 Spike (S) glycoprotein (S1, 158 peptides and S2, 157 peptides) in the presence of 1□μg/ml of costimulatory monoclonal antibodies to CD28 and CD49d. Samples mock-stimulated with phosphate□buffered saline (PBS)/dimethyl sulfoxide and costimulatory antibodies served as negative controls. Positive (phytohemagglutinin) and isotype controls were run in parallel. Cells were analyzed on a FACSCanto flow cytometer using DIVA v8 software (BD Biosciences Immunocytometry Systems, San Jose, CA).

**Supplementary Figure 2**. SARS-CoV-2-S plasma antibody levels as measured by Roche Elecsys® Anti-SARS-CoV-2 S immunoassay in nursing home residents (NHR) with or without comorbidities following full dose vaccination. The *P*-value for comparison is shown.

