## Supplementary figures and images for "B and T cell immune responses elicited by the BNT162b2 (Pfizer–BioNTech) COVID-19 vaccine in nursing home residents"

### Supplementary Figure 1

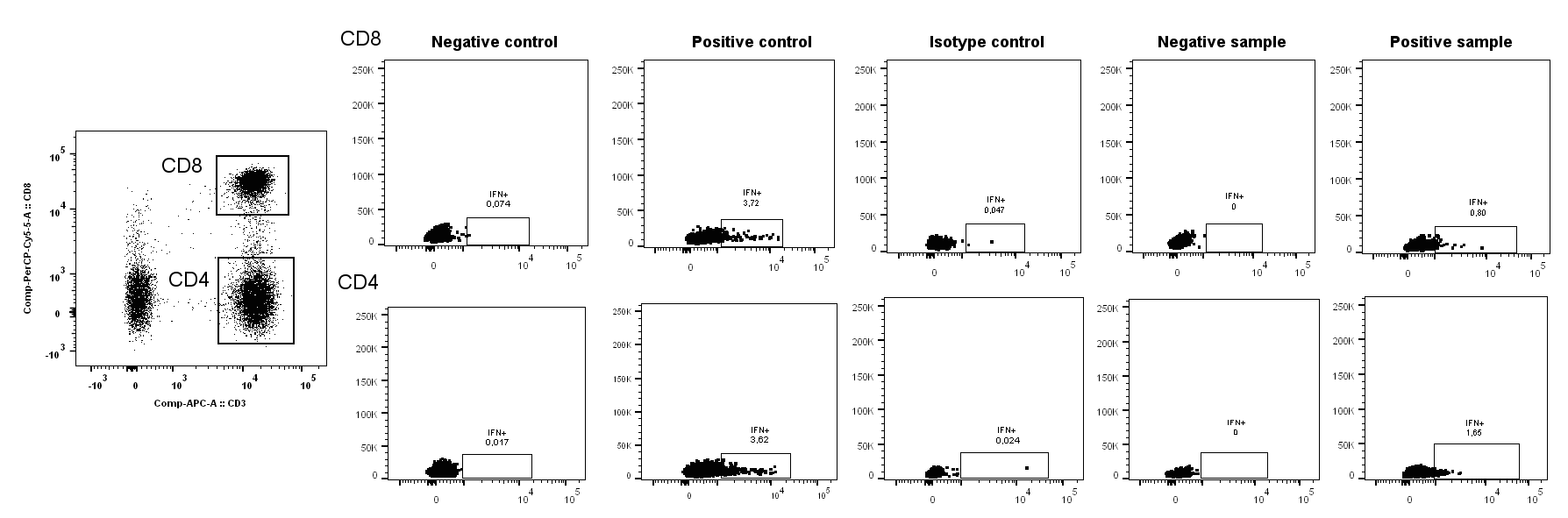

### Supplementary Figure 2

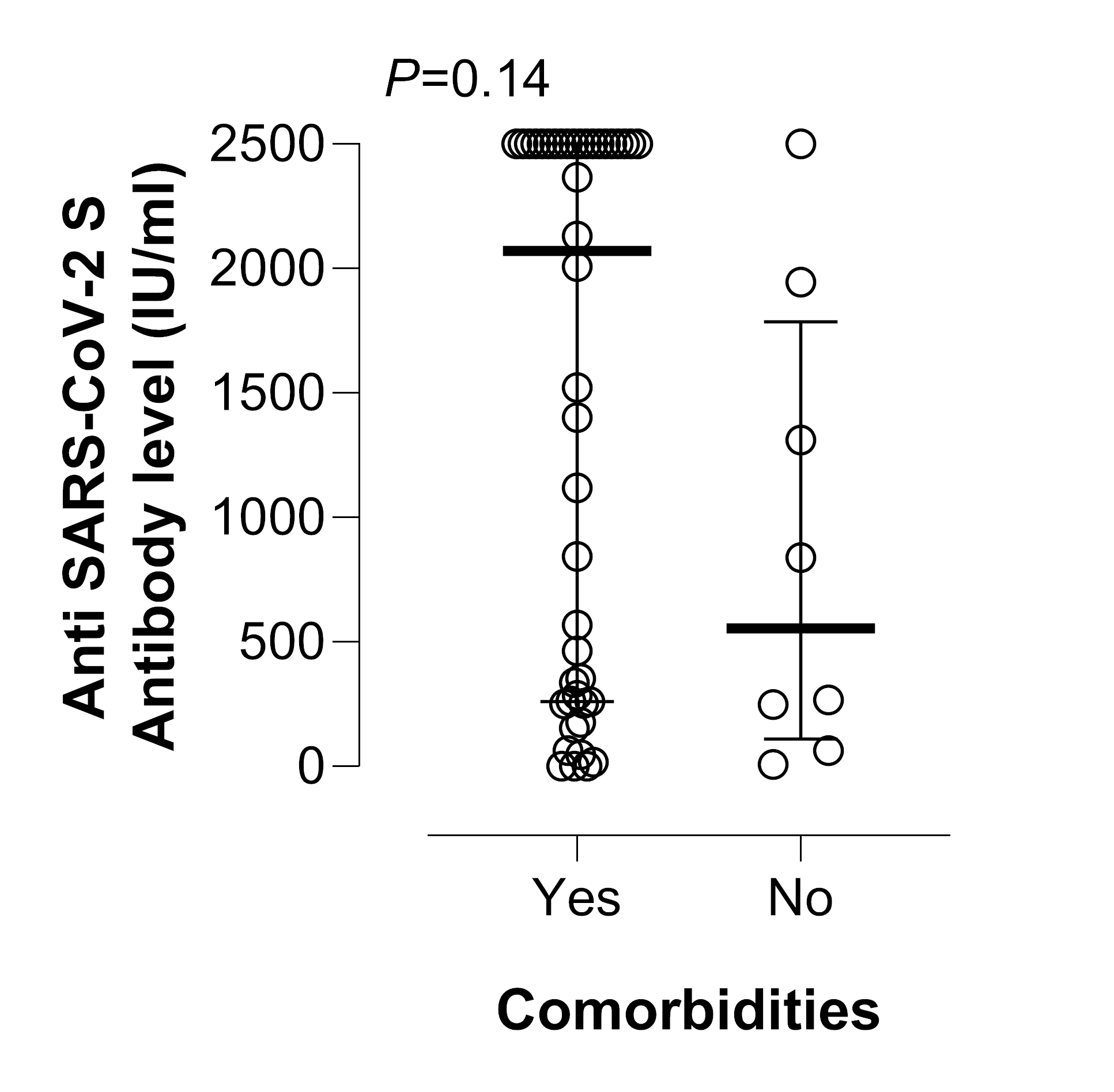
